# Generative AI as a Tool for Environmental Health Research Translation

**DOI:** 10.1101/2023.02.14.23285938

**Authors:** Lauren B. Anderson, Dhiraj Kanneganti, Mary Bentley Houk, Rochelle H. Holm, Ted Smith

**Affiliations:** Christina Lee Brown Envirome Institute, School of Medicine, University of Louisville, Louisville, KY 40202, United States; Department of Urban and Public Affairs, College of Arts and Sciences, University of Louisville, Louisville KY 40208, United States; University of Louisville Superfund Research Center, Louisville, KY 40202, United States

**Author notes:** **Correspondence** Correspondence should be sent to Ted Smith, Superfund Research Center, School of Medicine, University of Louisville, 302 E Muhammad Ali Blvd, Louisville, KY 40202, United States.

## Abstract

Generative artificial intelligence, popularized by services like ChatGPT, has been the source of much recent popular attention for publishing health research. Another valuable application is in translating published research studies to readers in non-academic settings. These might include environmental justice communities, mainstream media outlets, and community science groups. Five recently published (2021-2022) open-access, peer-reviewed papers, authored by University of Louisville environmental health investigators and collaborators, were submitted to ChatGPT. The average rating of all summaries of all types across the five different studies ranged between 3 and 5, indicating good overall content quality. ChatGPT’s general summary request was consistently rated lower than all other summary types. Whereas higher ratings of 4 and 5 were assigned to the more synthetic, insight-oriented activities, such as the production of a plain language summaries suitable for an 8^th^ grade reading level and identifying the most important finding and real-world research applications. This is a case where artificial intelligence might help level the playing field, for example by creating accessible insights and enabling the large-scale production of high-quality plain language summaries which would truly bring open access to this scientific information. This possibility, combined with the increasing public policy trends encouraging and demanding free access for research supported with public funds, may alter the role journal publications play in communicating science in society. For the field of environmental health science, no-cost AI technology such as ChatGPT holds the promise to improve research translation, but it must continue to be improved (or improve itself) from its current capability.

## Introduction

Generative artificial intelligence (AI), popularized by services like ChatGPT, has been the source of much recent popular attention for publishing health research.^1,2^ The software can instantly create sophisticated content ranging from text-to-image to text-to-audio and video depending on user prompts.^3^ ChatGPT, released by OpenAI^4^ (San Francisco, CA, USA) in November of 2022, is a large language model trained on a vast amount of data generating human-like responses to queries and is often used in applications such as chatbots. Another valuable application is in translating published research studies to readers in non-academic settings. These might include environmental justice communities, mainstream media outlets, and community science groups. Given the number of challenges with writing peer-reviewed research and the follow-on cost and effort in additionally creating plain language summaries (PLS), generative AI may offer scientific researchers a valuable new tool for improving environmental health risk communication and access by the larger constituencies in society.

However, researchers must balance the potential benefits of generative AI, such as making science more equitable and increasing the diversity and accessibility of scientific knowledge, with potential risks, such as degrading the quality and trust in the research practice. Chatbots can produce convincing but inaccurate text, which might undermine the very trust research translation aims to build.^1,2^ The role of academic researchers in environmental health programs has been tied to not only methods but the essential working relationships for translation to public health action.^5^ Using an analytic rubric, we explored the use of ChatGPT as an alternative to traditional human expert summarization to support PLS as a form of research translation and provide suggestions for environmental health research translation.

## Methods

Five recently published (2021-2022) open-access, peer-reviewed papers, authored by University of Louisville environmental health investigators and collaborators, were submitted (Supplemental Material). The PLS were created by entering the full text of each paper into the ChatGPT interface (version February 4, 2023), followed by a series of instructions: “Summarize the findings of this study in 500 words,” “Summarize the research paper at an eighth-grade reading level,” “What is the most important finding of this study?” and “What are the real-world impacts of this study?”. The responses from ChatGPT were evaluated using a combination of Likert-scale, yes/no, and text to assess scientific accuracy, completeness, and readability at an 8^th^ grade level by one of the authors of each study. The author reviewers were blind to the use of the generative AI as the source of the summarization.

## Results and Discussion

The average rating of all summaries of all types across the five different studies ranged between 3 and 5, indicating good overall content quality. ChatGPT’s general summary request was consistently rated lower than all other summary types. Whereas higher ratings of 4 and 5 were assigned to the more synthetic, insight-oriented activities, such as the production of a PLS suitable for an 8^th^ grade reading level and identifying the most important finding and real-world research applications. The majority of these more insight-orientated summaries were judged acceptable for use with the public. Even across this limited collection of studies, there were a few authors who commented that the PLS was still too technical for general audiences. In some cases, ChatGPT’s attempt at simplification removed important detail – for instance, stating wellbeing was associated with cardiovascular diseases (CVD), rather than risk of CVD. Minor inaccuracies around study method interpretations were also observed. In one processed paper the ChatGPT PLS referenced questions about levels of pollution and greenness around the home when the study did not include these questions. Other research has reported ChatGPT created abstracts not always distinguishable from those written by a human expert thus bringing up the need for source disclosure, however medical discharge summaries with automation have been shown to still require manual checking by a human expert where an inaccuracy in a summary report may lead to patient safety issues.^2,6,7^

There are several critical research needs, especially around inclusion and environmental health justice research.^2^ This is a case where AI might help level the playing field, for example by creating accessible insights and enabling the large-scale production of high-quality PLS which would truly bring open access to this scientific information. This possibility, combined with the increasing public policy trends encouraging and demanding free access for research supported with public funds, may alter the role journal publications play in communicating science in society. Recommendations for future research include the need for rating by community members of paired PLSs and review of a larger number and wider variety of environmental health research study types. For the field of environmental health science, no-cost AI technology such as ChatGPT holds the promise to improve research translation, but it must continue to be improved (or improve itself) from its current capability.

## Data Availability

The list of peer-reviewed research papers analyzed during the current study are available in the supplement to this manuscript.

## Acknowledgments

We would like to acknowledge the support of the Superfund Research Center at the University of Louisville (NIEHS Award Number P42ES023716). Special thanks are extended to the authors who reviewed and scored the summaries.

## Supplemental Material

List of peer-reviewed research papers authored by University of Louisville Envirome Institute environmental health investigators and collaborating research partners entered into the ChatGPT interface:

---

**Table S1.**
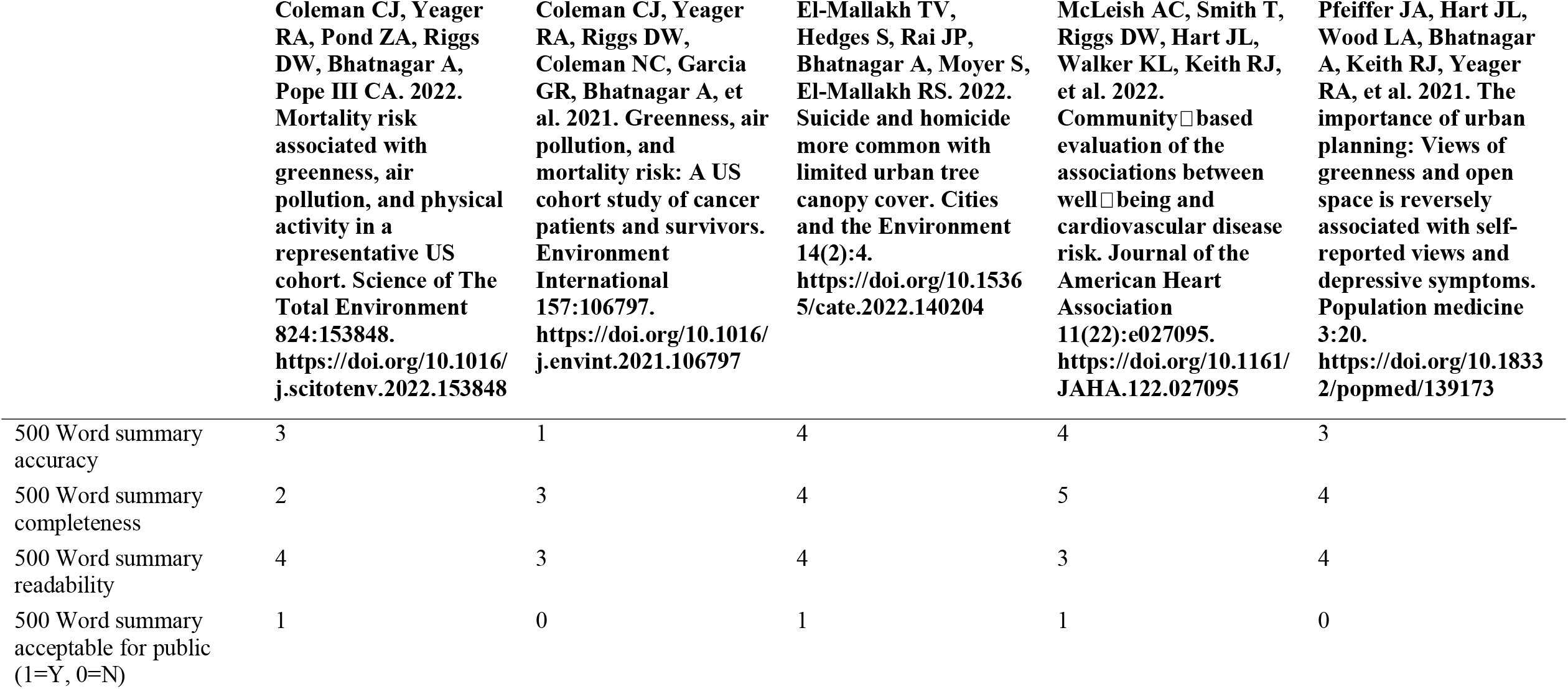
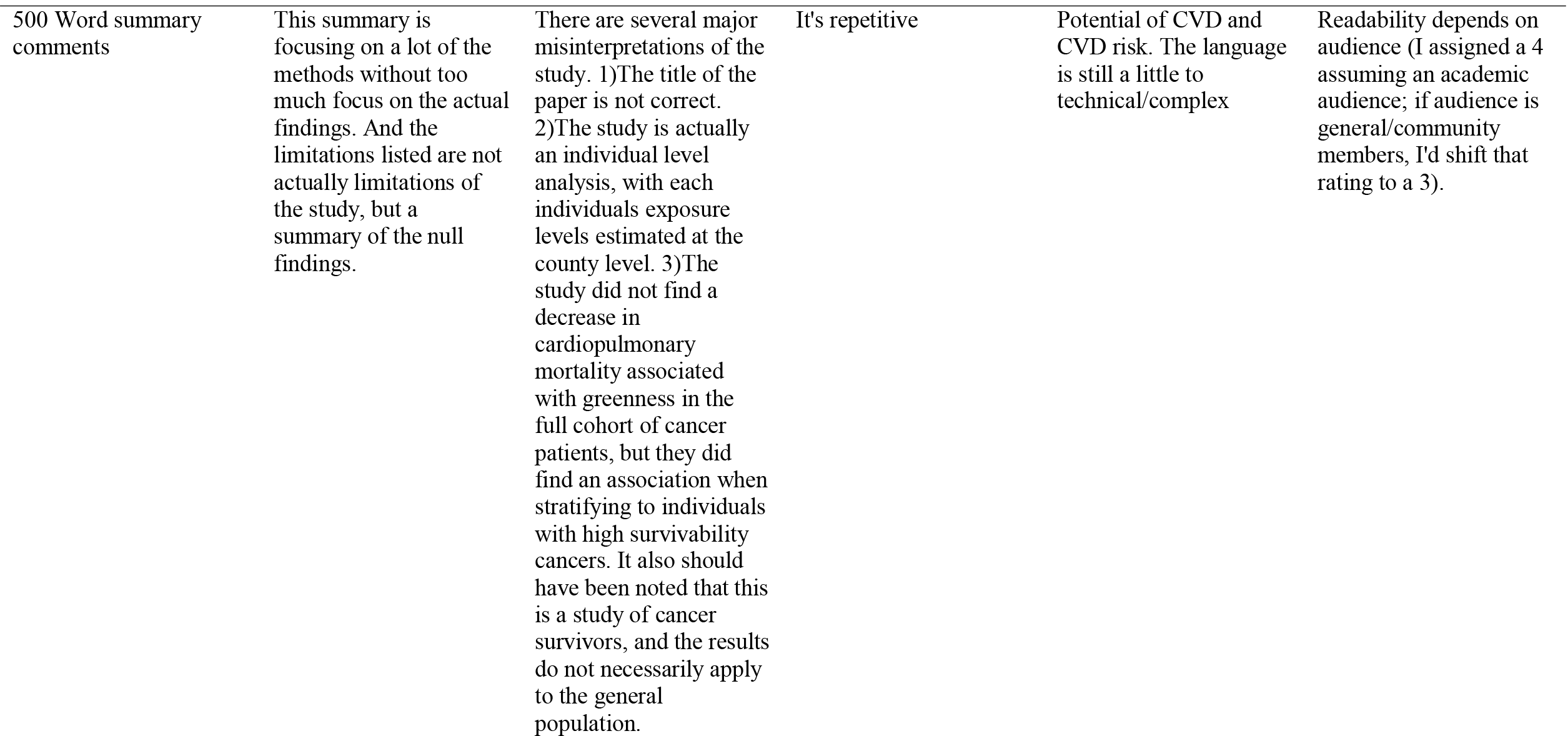
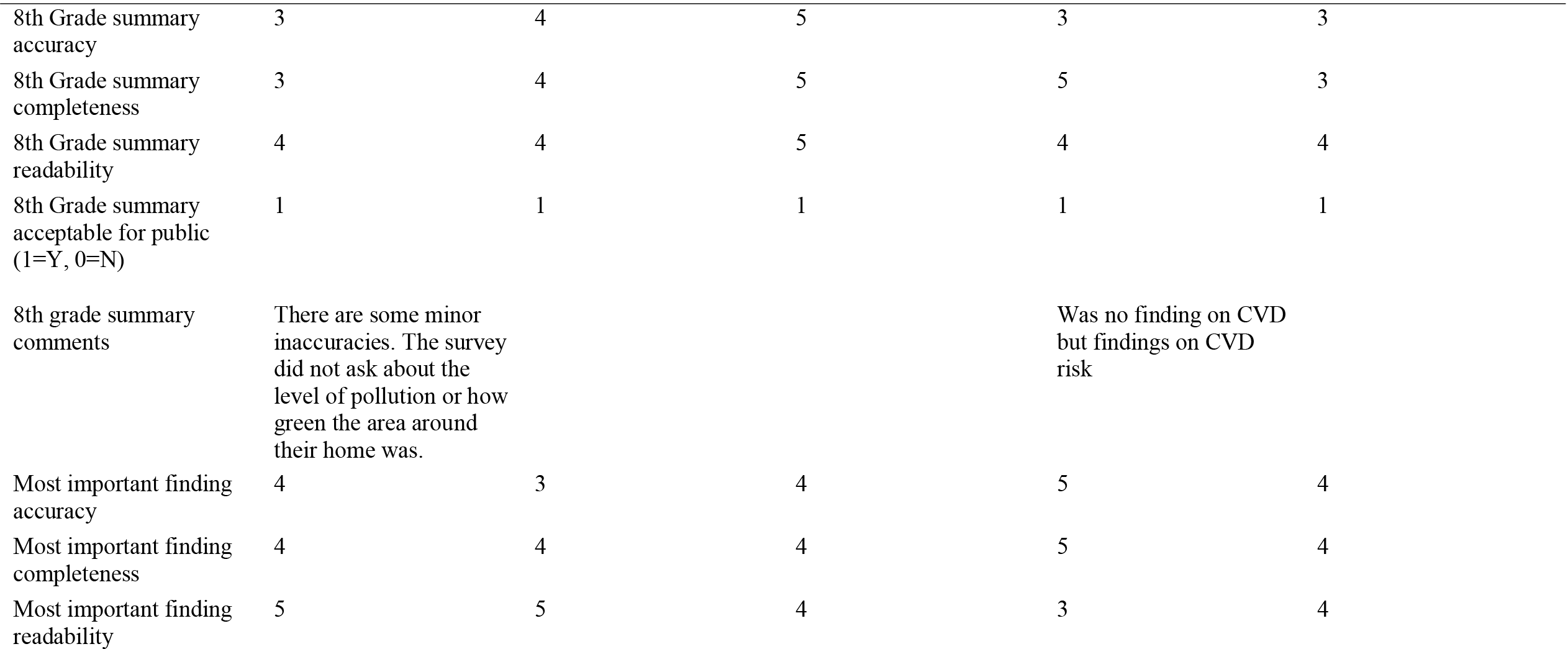
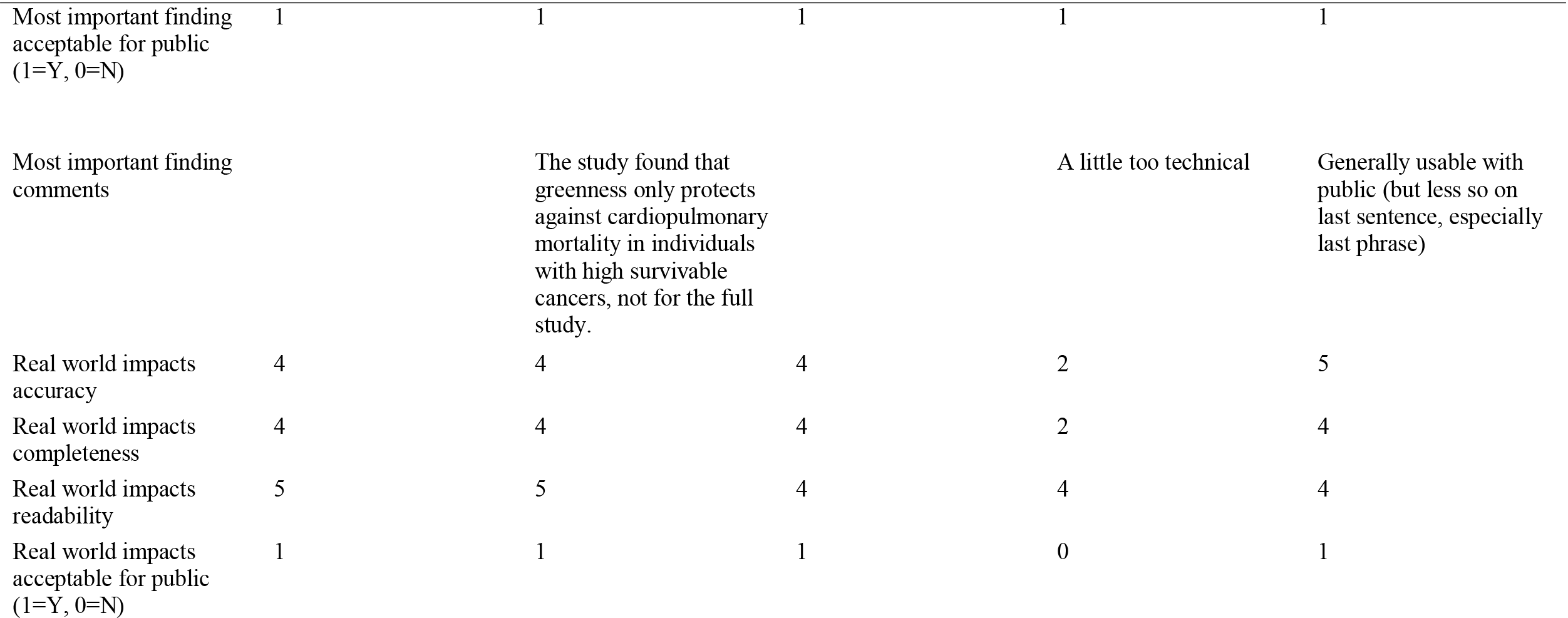
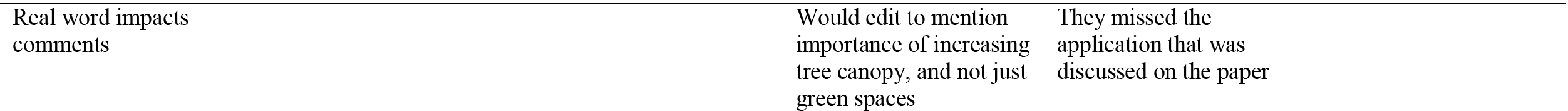
Evaluation responses from studied peer-reviewed papers

